# Efficacy and safety of *Andrographis paniculata* extract in patients with mild COVID-19: A randomized controlled trial

**DOI:** 10.1101/2021.07.08.21259912

**Authors:** Kulthanit Wanaratna, Pornvimol Leethong, Nitapha Inchai, Wararath Chueawiang, Pantitra Sriraksa, Anutida Tabmee, Sayomporn Sirinavin

## Abstract

**Objectives:** To assess the efficacy and safety of *Andrographis paniculata* extract (APE) in adults with mild COVID-19.

**Methods:** Sixty-three adults aged 18-60 years, without co-morbidity, with laboratory-confirmed mild COVID-19, were randomized 1:1 to receive APE (60 mg andrographolide, t.i.d, for 5 days) or placebo within 24 hours after admission, plus standard supportive care. The outcomes were clinical recovery rates by Day 5 using self-assessment scores, pneumonia by chest X-rays, nasopharyngeal SARS-CoV-2 detection by rRT-PCR on Day 5, changes of serum CRP levels, and adverse drug reactions. Chest X-rays and blood tests for CRP, liver and renal function, were performed on Days 1, 3, and 5.

**Results:** Baseline characteristics of patients in the APE-treatment (n=29) and placebo-control (n=28) groups were comparable. None had self-assessment scores showing complete clinical recovery by Day 5. Pneumonia occurred in 0/29 (0%) versus 3/28 (10.7%), (p=0.112). On Day 5, patients with SARS-CoV-2 detection were 10/29 (34.5%) versus 16/28 (57.1%), (p=0.086); patients with CRP >10 mg/L were 0/29 (0%) versus 5/28 (17.9%), (p=0.023), for APE-treatment and placebo-control groups, respectively. All three patients with pneumonia had substantially rising serum CRP; and high CRP levels on Day 5. None had evidence of hematologic, hepatic or renal impairment.

**Conclusion:** Even though the study was limited by small sample size, our findings suggested promising efficacy and safety of the APE-treatment regimen in adults with mild COVID-19. Further studies, with adequate power to assure these findings, are required.

## Introduction

Coronavirus disease 2019 (COVID-19), caused by the novel severe acute respiratory syndrome coronavirus 2 (SARS-CoV-2), has become a global pandemic of the highest priority (World Health Organization, 2021). Clinical spectrum of the disease is categorized into asymptomatic, mild, moderate, severe, and critical illness. About 80% of cases are classified as asymptomatic and mild (fever or upper respiratory tract symptoms, malaise, headache, muscle pain, nausea, vomiting, diarrhea, loss of taste or smell, but without pneumonia), or moderate (pneumonia, with oxygen saturation ≥94% on room air). Mild COVID-19 may worsen to moderate and severe disease, while pneumonia is the primary indicator of more severe disease progression alongside multiple organ dysfunction or failure (Wu and McGoogan, 2020; National Institute of Health USA, 2021).

Predictors of a worse outcome in COVID-19 patients include host factors (e.g., gender, age, co-morbidities), disease characteristics (Wu and McGoogan, 2020; National Institute of Health USA, 2021), and laboratory parameters such as C-reactive protein (CRP), which is an inflammatory marker (Mueller et al., 2020; Potempa et al., 2020; Wang, 2020; Stringer et al., 2021). In normal healthy individuals, baseline CRP levels in the blood are reported to be less than 10 mg/L (World Health Organization, 2014; Wang, 2020). The main mechanisms of disease progression in COVID-19 are intracellular viral replication causing cell lysis or cell death, and hyperactivation of immune responses contribute to widespread hyperinflammation (van Eijk et al., 2021). In addition to symptomatic management of patients with COVID-19, a therapeutic drug that would limit the course of infection is urgently needed.

*Andrographis paniculata* (Burm.f.) Nees is an annual herb native to the Indian subcontinent and widely introduced, naturalized, and cultivated in Southeast Asia and China (Kew Science, 2019). It is one of the commonly used medicinal plants in Indian Ayurvedic medicine, Chinese and Thai traditional medicines, for the treatment of acute upper respiratory tract infection and diarrhea (National Drug Committee, Thailand 2000; The State Pharmacopoeia Commission of the People’s Republic of China, 2015; Banerjee et al., 2021). Its name in Thai is Fa Thalai Chon. The main constituent of *A. paniculata* is andrographolide which is a bioactive diterpene lactone. The therapeutic efficacy of *A. paniculata* is due to its various pharmacological activities, i.e., antiviral, anti-inflammatory, antipyretic, and immune regulation activities (Dai et al., 2019; Hossain et al, 2021). The adverse effects include headache, fatigue, allergic reactions, nausea, diarrhea, and mild hypotension, which are uncommon; while more severe adverse effects, although very rare, are urticaria or anaphylaxis (Thamlikitkul et al., 1991; Suriyo et al, 2017; Worakunphanich et al., 2021).

In silico studies revealed that andrographolide is an inhibitor of the main protease of SARS-CoV-2 which leads to prevention of viral replication (Murugan et al., 2020; Shi et al., 2020; Enmozhi et al., 2021; Hiremath et al., 2021). Furthermore, *in vitro* study also demonstrated that it acts on SARS-CoV-2 by preventing viral replication in cells, especially in late phase of viral life cycle. (Phumiamorn S, 2020; Sa-Ngiamsuntorn et al., 2021). The treatment effects of APE were explored in a small group (n=6) of mild COVID-19 volunteers; clinical improvement and SARS-CoV-2 reduction were detected.

Therefore, this randomized, double-blind, placebo-controlled trial was conducted to determine the clinical and virological efficacy, and safety of oral *A. paniculata* extract among adults with mildly symptomatic SARS-CoV-2 infection. Additionally, we explored the role of serum CRP in the prediction of disease progression in mild COVID-19.

## Patients and Methods

The study was conducted from December 2020 to March 2021, during the second COVID-19 epidemic wave in Thailand. During that period of time, anyone who tested positive for SARS-CoV-2, from nasopharyngeal swabs for real-time reverse transcription polymerase chain reaction (rRT-PCR) test, were promptly admitted to state quarantine hospitals. Samut Prakan Hospital and Nakhon Pathom Hospital, two state quarantine hospitals in central Thailand, were our study sites.

### Patients

The patient inclusion criteria were (1) age 18–60 years; with (2) diagnosis of SARS-CoV-2 infection by positive rRT-PCR test; (3) being admitted to the hospitals within 72 hours of illness; (4) presence of any signs and symptoms of mild COVID-19 (e.g., fever, cough, sore throat, malaise, headache, and diarrhea); and (5) not having signs and symptoms of pneumonia or abnormal chest imaging. Patients were excluded if they (1) had any underlying medical conditions that increased the risk of severe COVID-19; (2) had any conditions for which using *A. paniculata* should be avoided, i.e., history of allergy to *A. paniculata*, taking antihypertensive or anticoagulant drugs, pregnancy or breast-feeding; or (3) were receiving any antiviral drug. On admission to the study hospitals, the inclusion and exclusion criteria were verified. Patients were informed about the study project and those interested in study participation were requested to sign informed consent.

### Study treatment

The standardized alcoholic extract of the aerial part of *A. paniculata* (APE, AP extract) was used. Both APE and placebo capsules were produced by Thai Herbal Products Co., Ltd, Thailand, and both were [indistinguishable](https://www.bing.com/search?q=define+indistinguishable&FORM=DCTRQY). Each capsule of APE contains 20 mg of andrographolide. The patients received either oral APE or placebo, within 24 hours after admission (Day 1 of clinical trial), in addition to standard supportive care following the national clinical practice guideline. The APE-treatment patients received 3 capsules (60 mg. andrographolide) of APE while the placebo-control patients received 3 capsules of placebo, 3 times a day (t.i.d.), for 5 days.

### Outcome assessment

The primary outcomes were complete clinical recovery rates by Day 5. The secondary outcomes included pneumonia development during the course of illness, SARS-CoV-2 detection, serum CRP levels, and adverse drug reactions. Clinical recovery was self-assessed by the patients using visual analog scale (VAS) scores. Chest X-rays were assessed on Days 1, 3, and 5 of the clinical trial. Additional chest X-rays were performed for those without abnormal chest X-rays by Day 5, who had progression of respiratory symptoms. Nasopharyngeal swabs were performed on Day 5 for the detection of SARS-CoV-2 by rRT-PCR. Venous blood specimens were collected on Days 1, 3, and 5 for complete blood count, blood urea nitrogen and creatinine, aspartate aminotransferase and alanine aminotransferase, and CRP. Suspected clinical adverse effects of drugs were recorded.

### Sample size

Sample size was calculated from the primary outcome with assumption that the duration of symptoms in mild COVID-19 was about 5 days and that the AP-treatment would cause 2-day decrease in duration. This led to an estimated sample size of 26 participants per group to provide type I error of 5% and type II error of 20%. The intended sample size was adjusted to 32 participants per group to compensate for possible drop-out.

### Randomization and blinding

The 1:1 randomization was prepared by a central researcher before participant enrollment, by block randomization with a block size of six using a randomization software. The randomization codes were labelled on APE or placebo packages which were physically indistinguishable; each package contained 45 capsules of APE or placebo for 5-day treatment of individual patients. The study hospitals were provided with code-labelled packages of APE or placebo. Each new participant was assigned the code on the received drug package. The randomization code was kept secret from the clinic and the participating investigators, and the code was revealed at the end of the study for analysis.

### Statistical analysis

Data were described by mean with standard deviation (mean ± SD) or frequency (%), and compared between groups by Student t-test for continuous data, Chi-square test or Fisher exact test for categorical data. All analyses were performed by STATA version 16.0, and p-value <0.05 was considered as being statistically significant.

## Results

Fifty-seven of the 63 enrolled patients completed the study, 29 patients in the APE-treatment group and 28 patients in the placebo-control group. Three patients of each group withdrew during the study, due to unwillingness for further blood testing (2 control, 2 APE-treatment) and unpleasant symptoms (1 palpitation in control, and 1 diarrhea in APE-treatment); while none of them developed pneumonia. The baseline characteristics (age, gender, body mass index, and CRP level on enrollment) were comparable between the 2 groups (Table1).

**Table 1.** Baseline characteristics of patients who received *Andrographis paniculata* extract (AP extract) or placebo

| Characteristics | AP extract<br>(n=29) | Placebo<br>(n=28) | *p*-value |
| --- | --- | --- | --- |
| Age (years), mean ± SD | 39.3 ± 11.4 | 39.4 ± 11.6 | 0.949 |
| Male, n (%) | 12(41.4) | 11 (39.3) | 0.870 |
| BMI (kg/m<sup>2</sup>), n (%) |  |  |  |
| Underweight or normal weight | 19 (65.5) | 15 (53.6) | 0.684 |
| Overweight (BMI 25-29.9 kg/m<sup>2</sup>) | 7 (24.1) | 9 (32.1) |  |
| Moderate Obesity (BMI ≥ 30-34.9 kg/m<sup>2</sup>) | 3 (10.3) | 4 (14.3) | |
| Severe Obesity (BMI ≥ 35 kg/m<sup>2</sup>) | 0 (0.0) | 0 (0.0) | |
| CRP levels on hospital admission, n (%) |  |  |  |
| ≤5 mg/L | 24 (82.8) | 22 (78.6) | 0.495 |
| >5-10 mg/L | 5 (17.2) | 4 (14.3) |  |
| >10-18 mg/L | 0 (0.0) | 2* (7.1) |  |

BMI = body mass index, CRP = C-reactive protein
\*One patient with rising CRP levels developed pneumonia, the other with steady CRP levels did not.

None of the participants had complete clinical recovery by Day 5 as determined by patient self-assessed VAS scores, and further assessment was not performed according to the protocol. Table 2 showed the numbers of patients with pneumonia, Day-5 detection of nasopharyngeal SARS-CoV-2, and CRP levels >10 mg/L in the two groups. None of the patients in the APE-treatment group, but 3 patients in the placebo-control group developed pneumonia (on Days 6-7), being 0/29 (0%) versus 3/28 (10.7%), (p=0.112). On Day 5, SARS-CoV-2 was detected from nasopharyngeal swab in 10/29 (34.5%) patients of the APE-treatment group, and 16/28 (57.1%) patients of the placebo-control group, (p=0.086).

**Table 2.**
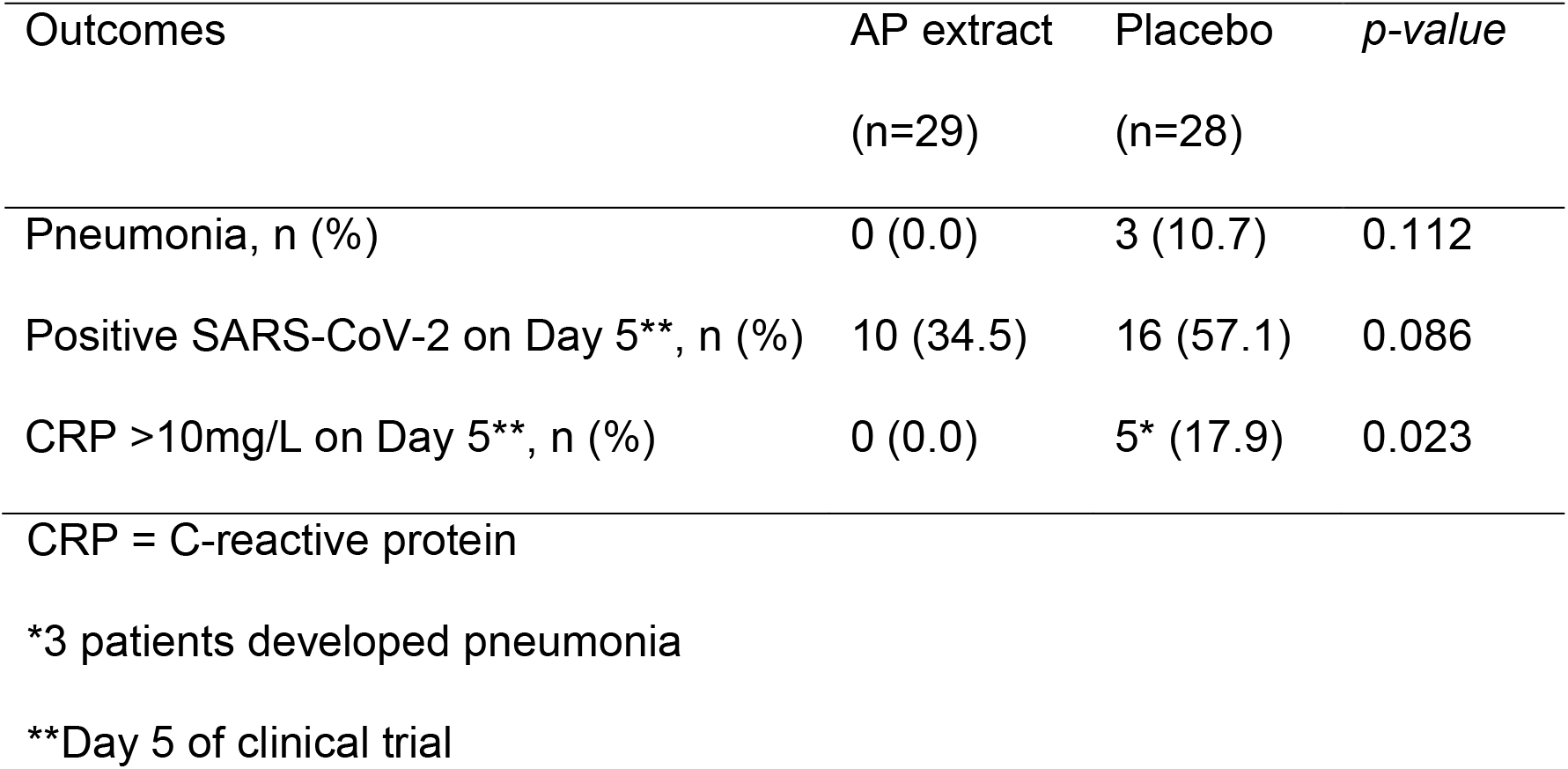
The study outcomes in the groups of patients who received *Andrographis paniculata* extract (AP extract) or placebo.

| Outcomes | AP extract<br>(n=29) | Placebo<br>(n=28) | *p-value* |
| --- | --- | --- | --- |
| Pneumonia, n (%) | 0 (0.0) | 3 (10.7) | 0.112 |
| Positive SARS-CoV-2 on Day 5**, n (%) | 10 (34.5) | 16 (57.1) | 0.086 |
| CRP >10mg/L on Day 5**, n (%) | 0 (0.0) | 5* (17.9) | 0.023 |

CRP = C-reactive protein
\*3 patients developed pneumonia
\*\*Day 5 of clinical trial

None of the APE-treatment group had CRP >10 mg/L during the 5-day study period. On admission, 2 patients in the placebo-control group had high CRP levels (18.3 and 18.4 mg/L), and one who had rising CRP levels developed pneumonia while another with steady CPR levels did not. On Day 5, the proportions of patients with CRP >10 mg/L were 0/29 (0%) in APE-treatment group, and 5/28 (17.9%) in placebo-control group, (p=0.023). Three patients who had pneumonia had substantially rising serum CRP on Day 3 and high CRP of 13.9, 20.7, and 21.0 mg/L on Day 5. In addition, 2 patients in the placebo-control group had rising CRP and high CRP of 16.0 and 16.1 mg/L on Day 5, but did not have pneumonia.

Abnormal symptoms as possible AP adverse effects were found in 2 patients in the APE-treatment group, and both were mild and self-limited gastrointestinal tract symptoms (1 diarrhea), and one patient in the placebo-control group had palpitation. None of the patients in both groups had laboratory evidence of hematologic, hepatic or renal function impairment.

## Discussion

The study of the efficacy of APE-treatment on the primary outcome, as clinical recovery rates by Day 5, could not give relevant information. However, our study suggests that the regimen of APE treatment (oral 60 mg of andrographolide, t.i.d., for 5 days) given early to adults with mild COVID-19, without risk for severe COVID-19 by host factors, has promising efficacy on mild COVID-19, though the sample size is too small to detect statistically significant difference of outcomes. The study suggests a tendency of risk reduction of pneumonia, more rapid SARS-CoV-2 clearance, and inflammation suppression. The potential treatment benefit of APE on COVID-19 is likely to be the result of the anti-SARS-CoV-2 and anti-inflammatory activities of andrographolide and its derivatives. Adverse effects from the treatment were found uncommon and benign.

A recent comprehensive review on antimicrobial pharmacology, clinical safety, and efficacy of *Andrographis paniculata*, demonstrated its high potential for effective and safe COVID-19 treatment, and there was a need for clinical evidence (Hossain et al., 2021). This study was performed on orally-administered APE in mild COVID-19 patients. A recent study from China on injectable andrographolide (Xiyanping in Chinese) in mild to moderate COVID-19 patients, demonstrated significant reduction of the time to cough relief, fever resolution and virus clearance, and less patients experienced disease progression to the severe stage. (Zhang et al., 2021).

The second epidemic wave of SARS-CoV-2 in Thailand lasted between December 2020 and March 2021, the duration which covered the study period. Using the whole genome sequencing, it was found that approximately 90% of the strains, circulating in the study area during that period, belonged to the B.1.36.16 lineage (COVID-19 Network Investigations Alliance, 2021). Considering that the treatment effects of andrographolide may vary with different SARS-CoV-2 strains, further exploration is therefore needed.

Three patients who had pneumonia had substantially rising serum CRP on Day 3 and high CRP of 13.9, 20.7, and 21.0 mg/L on Day 5. These findings of this study on serum CRP levels in mild COVID-19 patients agree with the previous retrospective study in 27 adults in the early stage of COVID-19 that CRP levels were low in mild disease and increased with disease progression (Wang, 2020). A study reported that increasing CRP levels during the first 48-to-72 hours of hospitalization was a better predictor of respiratory deterioration than initial CRP levels, while steady CRP levels were observed in patients whose condition remained stable (Mueller et al., 2020).

The proportions of the 3 secondary outcomes (pneumonia, viral detection, CRP) in the 2 compared groups of this trial suggested benefit of APE-treatment in mild COVID-19. A statistically non-significant difference (p ≥0.05) in this underpowered study does not imply that the treatment is not clinically effective (Tsang R, 2009).

## Conclusions

This double-blind RCT, in adults with mild COVID-19 and without risk factors for severe disease, suggested promising effects of the oral APE-treatment regimen on the prevention of pneumonia, shortening of viral shedding period, and inflammatory suppression. Adverse drug reactions were mild, self-limited, and uncommon. The study was limited by the small sample size. Further studies, with adequate power to assure conclusions, are required.

## Data Availability

Manuscript Number: THEIJID-D-21-01999
Article Title: Efficacy and safety of Andrographis paniculata extract in patients with mild COVID-19: A randomized controlled trial.
International Journal of Infectious Diseases.
On Peer review process.

## Acknowledgments

The authors gratefully acknowledge the study team, at Samut Prakan Hospital and Nakhon Pathom Hospital, who involved in this study; the Department of Medical Science, Ministry of Public Health (MOPH), for virological study; Dr. Marut Jirasetsiri and Dr. Amporn Benjaponpitak, Department of Thai traditional and Alternative Medicine, MOPH, for their full encouragement and support. We would like to thank Stephen John Pinder and Dr. Anchalee Chuthaputti for conducting a language review on this manuscript.

## Ethics approval and consent to participate

Ethical approval was obtained from the Ethics Committee for Research in Human Subjects in the Fields of Thai Traditional and Alternative Medicine (No.12-2563). All participants gave written informed consent.

